# ChatGPT as a decision-support tool for better self-monitoring of hearing

**DOI:** 10.1101/2024.12.16.24319073

**Authors:** Małgorzata Pastucha, Anna Ratuszniak, Małgorzata Ganc, Edyta Piłka, Iryna Drohobycka, Henryk Skarżyński, W. Wiktor Jedrzejczak

## Abstract

**Background:** The rapid development of large language model chatbots, such as ChatGPT, has created new possibilities for healthcare support. This study investigates the feasibility of integrating self-monitoring of hearing (via a mobile app) with ChatGPT’s decision-making capabilities to assess whether specialist consultation is required. In particular, the study evaluated how ChatGPT’s accuracy to make a recommendation changed over periods of up to 12 months.

**Methods:** ChatGPT-4.0 was tested on a dataset of 1,000 simulated cases, each containing monthly hearing threshold measurements over periods of up to 12 months. Its recommendations were compared to the opinions of 5 experts using percent agreement and Cohen’s Kappa. A multiple-response strategy, selecting the most frequent recommendation from 5 trials, was also analyzed.

**Results:** ChatGPT aligned strongly with the experts’ judgments, with agreement scores ranging from 0.80 to 0.84. Accuracy scores improved to 0.87 when the multiple-query strategy was employed. In those cases where all 5 experts unanimously agreed, ChatGPT achieved a near-perfect agreement score of 0.99. It adapted its decision-making criteria with extended observation periods, seemingly accounting for potential random fluctuations in hearing thresholds.

**Conclusions:** ChatGPT has significant potential as a decision-support tool for monitoring hearing, able to match expert recommendations and adapting effectively to time-series data. Existing hearing self-testing apps lack capabilities for tracking and evaluating changes over time; integrating ChatGPT could fill this gap. While not without its limitations, ChatGPT offers a promising complement to self-monitoring. It can enhance decision-making processes and potentially encourage patients to seek clinical expertise when needed.

**Graphical abstract:** 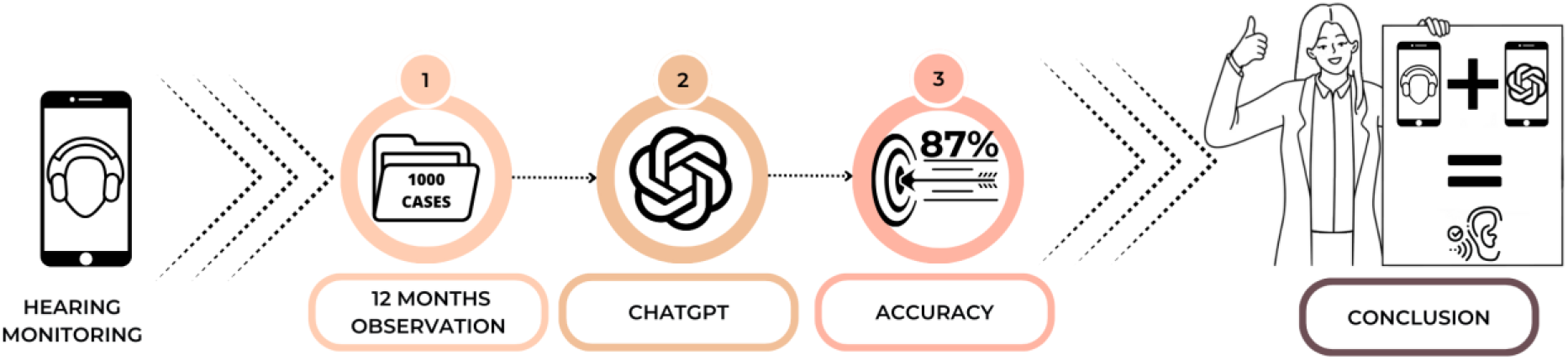

## 1. Background

Hearing loss is an increasing global health concern [1], driven by factors such as an aging population and widespread exposure to ototoxic substances and excessive noise [2–4] [5–7] [8]. Research on high-risk groups (e.g. chemotherapy patients) highlights the critical need for continuous monitoring of hearing thresholds [9,10]. However, nearly everyone faces some degree of risk for hearing loss, making the issue more universal than it first seems. Early detection of changes in hearing is important for preventing further deterioration and mitigating the long-term effects of deafness, which can impact communication, cognitive health, and quality of life [11–13]. Expanding access to advanced hearing monitoring technologies offers a promising option, empowering individuals to take proactive steps in preserving their auditory health.

Despite considerable advances in screening tools, effective monitoring of hearing remains a significant challenge [14,10]. The outcome of a single test does not always provide sufficient information to decide whether there has been a significant decline in hearing. This is particularly the case where there are only minor changes [15], irregular fluctuations [16], or slow decline of hearing [17]. To enhance the efficacy of change detection, it might be better to systematically monitor hearing over longer periods of time (even monthly over a year), which would enable trend analysis of measurements.

But such an approach involves frequent visits to an audiologist, which is hardly practical. A more realistic approach may be use a mobile app to self-test hearing at home, and in fact mobile hearing apps are becoming increasingly popular [18]. Home testing is convenient and eliminates the need for frequent visits to a specialist [19,20]. The results of recent studies indicate that there is a high degree of concordance between app-based readings and those performed clinically [10,21].

Notwithstanding the potential of apps, there is a lack of clear criteria for interpreting the results. How bad does my hearing have to be before I consult a specialist? [22]. A variety of assessment methods can be found in the literature [23–25], a situation that makes it difficult to interpret the results and hence the opportunities for early intervention. It is obviously impossible for people to self-test their hearing each month and then ask an audiologist for an opinion – there aren’t enough audiologists for that.

In this context, artificial intelligence (AI)-based systems, including chatbots based on large language models (LLMs) such as ChatGPT, might fill the gap. These models could analyze complex data patterns [26,27] and make recommendations on the need for specialist consultation, based on the number and nature of changes shown in self-reported measurements. This would provide patients with more personalized support, thereby motivating them to monitor their hearing regularly.

Despite the growing role of AI and LLMs in fields such as audiology [28–32] and otolaryngology [33,34], and some promising applications in clinical decision-making [35], there is a lack of studies assessing whether such systems can reliably support decision-making in a hearing monitoring context.

The objective of the present study was to assess how well ChatGPT could perform in helping to make decisions about whether a hearing specialist should be consulted based on a series of smartphone hearing tests. In particular, we wanted to know, first, whether ChatGPT’s recommendations aligned with what human experts would recommend; second, whether ChatGPT’s decisions were simply based on rigid, predefined criteria or were more nuanced than that, i.e. whether ChatGPT would adjust its recommendation criteria when provided with more extensive data over a longer observation period.

## 2. Material and methods

The study involved two main parts. For the first, we compared ChatGPT decisions with experts and rigid criteria. And in the second part of the study we determined the threshold for decision for different monitoring lengths. Details of the methods used and the testing procedure in each part of this study are presented below.

### 2.1. First part – Comparison of ChatGPT decisions with experts and rigid criteria

#### 2.1.1. Study procedure

This part involved several steps. For the first, a synthetic database of average hearing thresholds was created, reflecting hearing monitoring with 12 measurements (i.e. once every month over a year). This data was delivered to ChatGPT 4o (OpenAI, USA) with a question about whether a chosen test subject should visit a specialist. Next, the agreement between ChatGPT and 5 audiological experts was assessed. They were also compared with a rigid cut-off criterion of a change of 10 dB in hearing acuity.

A schematic of the study is shown in Figure 1, and the individual steps are described in more detail in the following.

**Figure 1.**
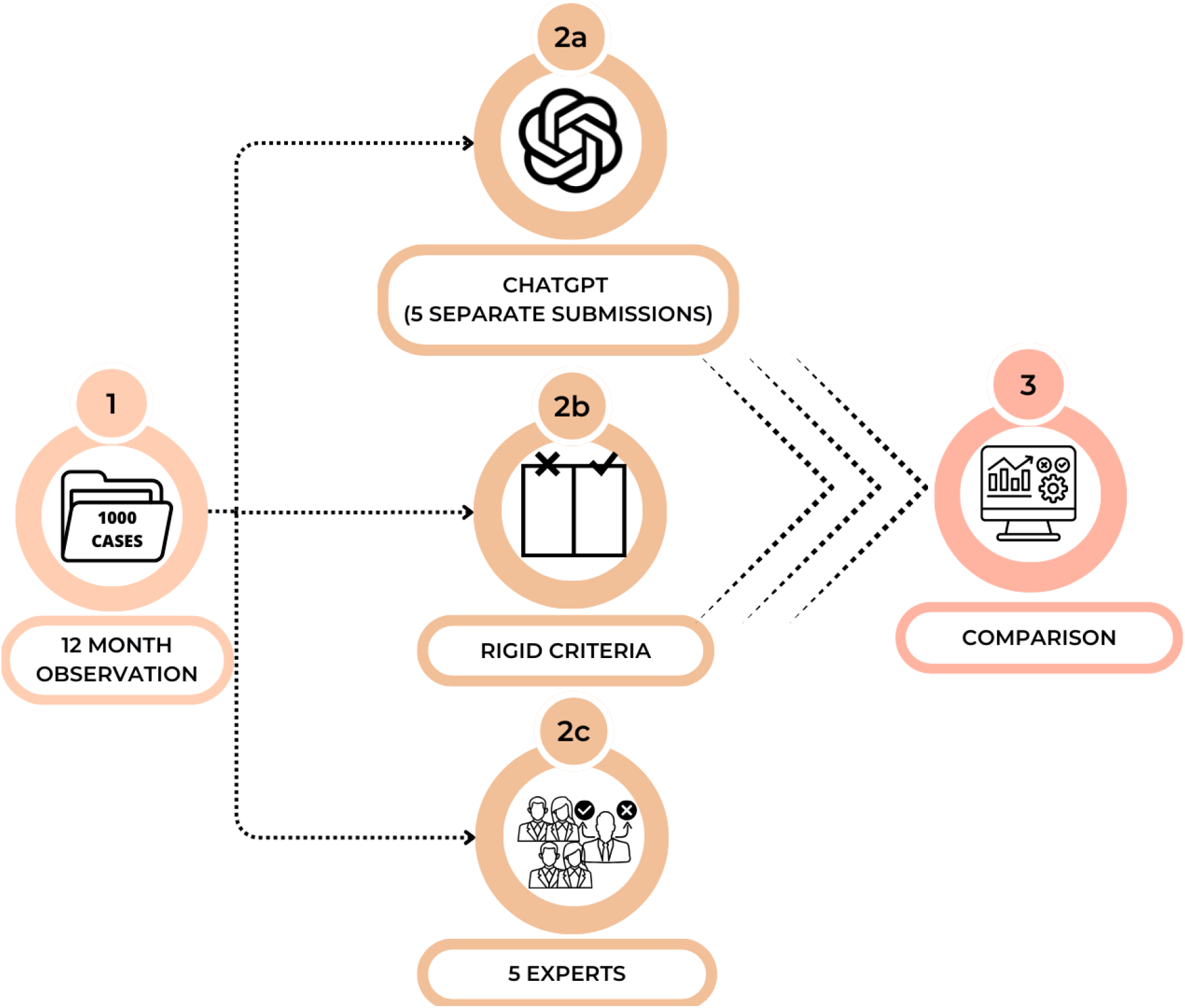
Outline of the study’s procedures for the first part of the study

#### 2.1.2. Data set

##### 2.1.2.1. Simulated data

Because of limited access to long-term hearing data and the necessity to safeguard sensitive data, the study used simulated data instead. Two experts developed the cases by first looking at actual patient data and identifying typical fluctuations in hearing thresholds over time. Based on their knowledge, experience, and the available literature [36,37], the experts constructed a synthetic database of 1,000 cases reflecting diverse scenarios in which hearing thresholds changed over time.

Each case contained a reference point (corresponding to a clinical examination) followed by self-monitoring data for 12 months (corresponding to tests made using a mobile app). The pure tone average (PTA) was taken as the representative variable. PTA is the average of 0.5, 1, 2, and 4 kHz, and is often used in many hearing test apps [38–41].

##### 2.1.2.2. Creation of database and examples

The database containing the reference measurement and 12 months of follow-up was created. Cases were created on the basis of 3 main criteria: initial degree of hearing loss (5 possibilities), level of change in threshold in consecutive measurements (0 to 39 dB in 1 dB steps), and type of change (another 5 possibilities). In this way, there were 5 × 40 × 5 = 1000 combinations. Figure 2 schematically shows the factors employed to construct the 1000 cases.

**Figure 2.**
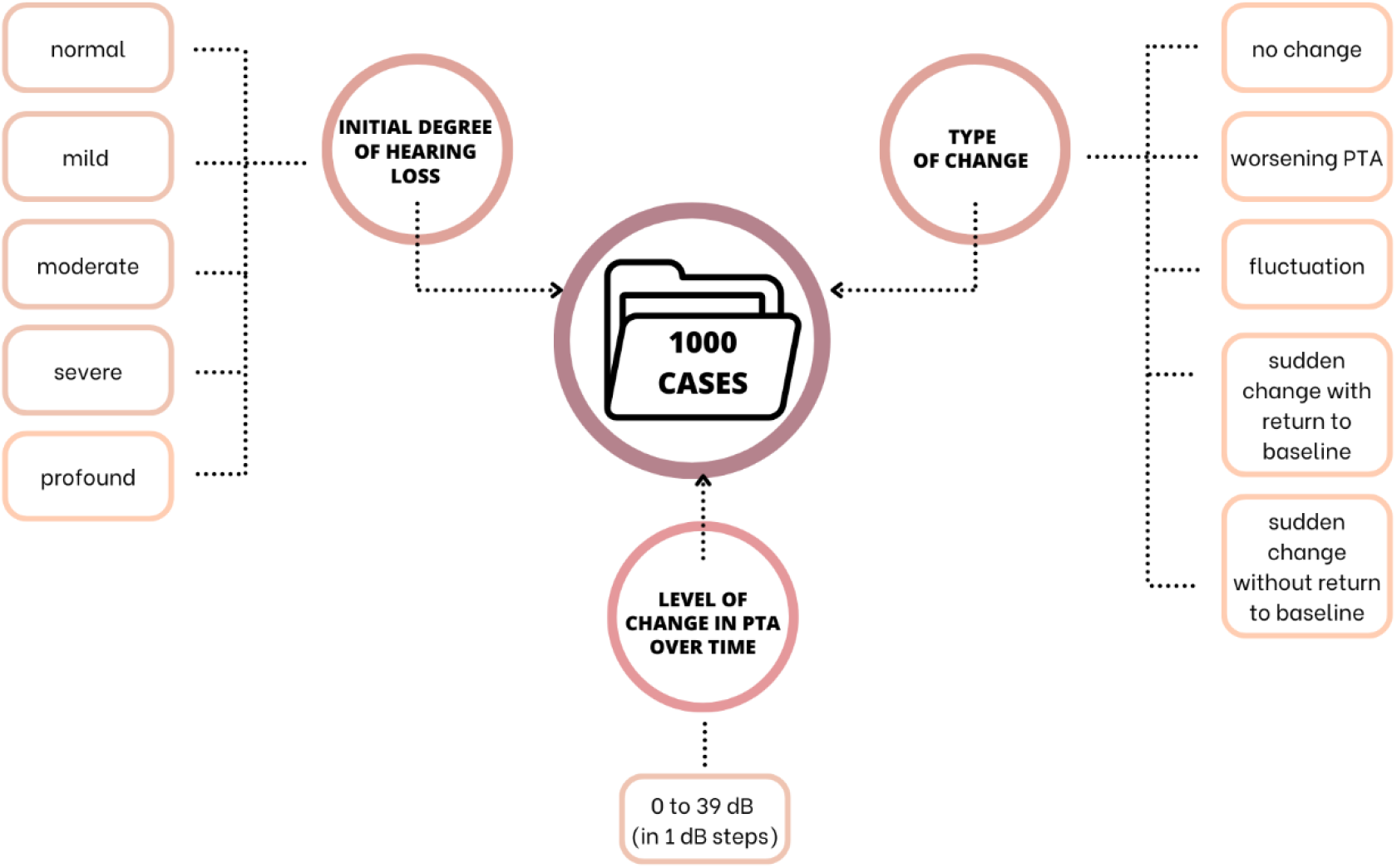
Schematic representation of information used to create each case.

In more detail, cases were assigned to one of 5 categories of hearing loss: normal, mild, moderate, severe, and profound in accordance with World Health Organization (WHO) guidelines [42]. A change in PTA over time (0 to 39 dB in 1 dB steps) was assigned to each category. A maximum of 39 dB was used on the basis that such a significant change in hearing would be obvious to the patient and so they would no doubt contact a specialist even without self-testing. Finally, 5 types of change were possible: no change (stable), worsening PTA, fluctuating over time, a sudden change with return to baseline, and a sudden change without return to baseline. Examples of cases representing the 5 types of changes are illustrated in Figure 3.

**Figure 3.**
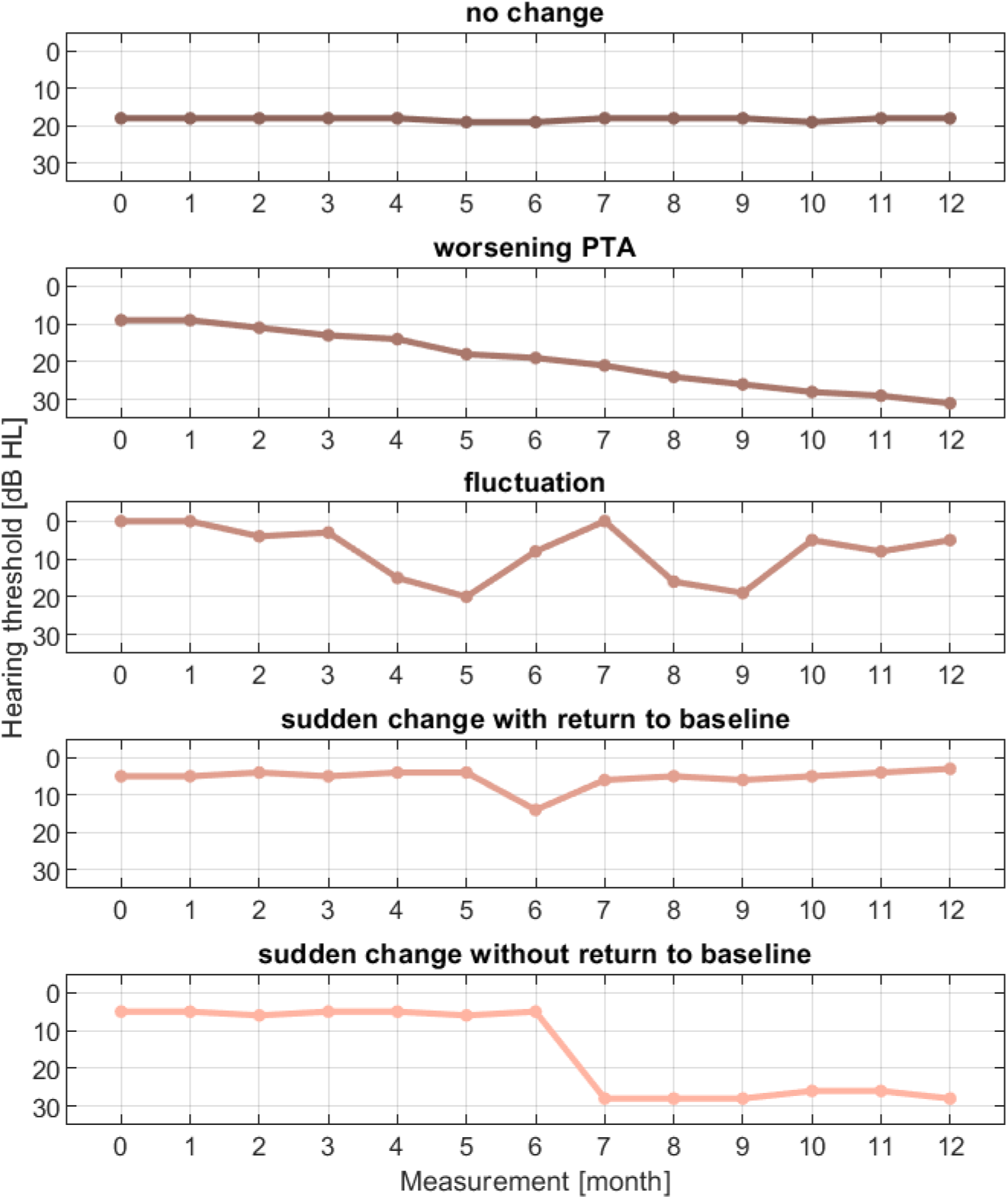
Examples of synthetic data from a pool of 1,000 hypothetical cases. The dimension illustrated here represents the 5 different types of changes in PTA typically encountered in a clinic (as indicated in the chart titles). The y-axis represents Pure Tone Average (PTA), while the x-axis indicates measurement periods in months. Month 0 corresponds to the initial test at the clinic (baseline), while months 1–12 reflect self-tests conducted by each patient.

In the end, the cases were therefore made up of a combination of 5 × 5 × 40 categories, thus allowing the creation of a database of 1000 cases covering just about every possible hearing threshold change. The complete dataset, comprising 1000 cases, can be found in the supplementary material

#### 2.1.3. Provision of the dataset to ChatGPT

Our dataset reflected the following scenario. A patient is tested by an audiologist in the clinic and then continues self-testing their hearing for some period (obtaining a set of PTAs). Then the patient (or a clinician who is monitoring the patient’s data) asks ChatGPT if, based on the data, they should consult an audiologist for a professional opinion.

In this study, a version of ChatGPT-4o (OpenAI, USA) was utilized. The LLM was provided with data in the form of CSV files, with each file containing the synthetic hearing thresholds for 50 patients. (Because of limitations to the amount of data ChatGPT could accept, we made 20 submissions each of 50 patients, producing 1000 cases.) The instructions provided to ChatGPT were as follows:,, *The average hearing threshold was measured in 50 patients at the clinic. Each patient then had their hearing threshold measured every month for the last twelve months using a mobile app. I attach a table with all the results. The first column (no) contains the patient number. The second column (clinic) contains the average thresholds measured at the clinic, and the following columns (app_1, 2*…*) contain the thresholds measured with the mobile app in the following months. Based on these data, decide whether a patient should visit a specialist. For each patient, give the answer: No (0); or Yes (1)”*.

The queries for ChatGPT were provided by one of the researchers, and the responses were stored in a CSV file. Prior to the introduction of each new query, the session was refreshed in order to eliminate possible interactions with previous responses.

The entire procedure was repeated 5 times in case there were any variations in ChatGPT responses, but the instructions remained the same for all submissions.

#### 2.1.4. Experts

A group of 5 experts with several years of clinical experience in audiology were invited to participate and given identical instructions to those provided to ChatGPT. Each expert conducted an independent assessment based on their knowledge and experience and submitted their responses in the form of CSV files.

#### 2.1.5. Cut-off criterion

Currently, there are no clear guidelines specifying what change in PTA calls for a visit to a specialist. The existing recommendations [23,24] differ, making it difficult to settle on a single value. For the purposes of this study, a change in PTA of ≥10 dB was selected as the criterion indicating that a professional should be consulted. This choice was based on the methodology used in another hearing monitoring study [43] and a review of the literature on the repeatability of measurements made using automated audiometers [44] and with mobile apps [45].

### 2.2. Second part – Determination of threshold for decision

#### 2.2.1. Study procedure

In the second part of the study, a synthetic database of average hearing thresholds was created reflecting monthly different monitoring lengths, made up of 1, 2, 4, 8, or 12 measurements for each subset. As in the first part of the study, this data was delivered to ChatGPT 4o (OpenAI, USA) with a question about whether a chosen test subject should visit a specialist. The responses generated by ChatGPT for the 5 databases with different monitoring periods were analyzed to deduce if and how ChatGPT’s decision criteria changed as the monitoring length increased.

A schematic of the study procedure is shown in Figure 4, and the individual steps are described in more detail in the following.

**Figure 4.**
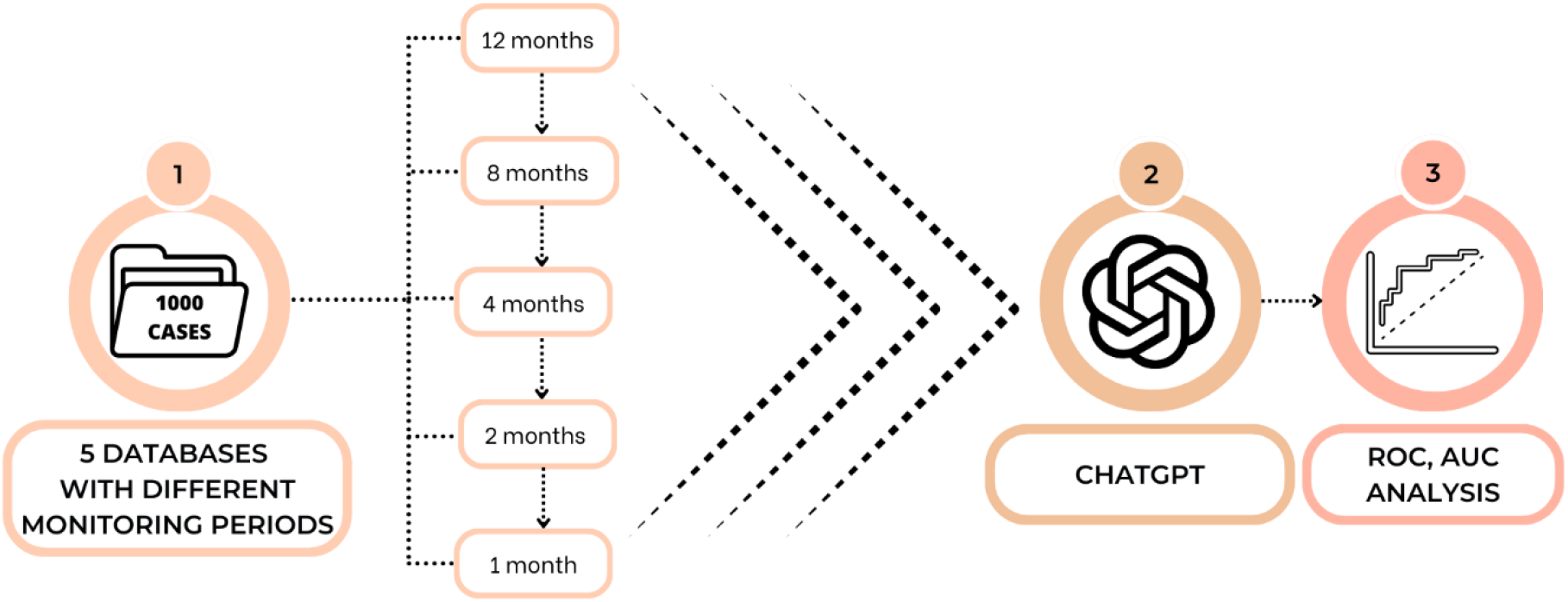
Outline of the study’s procedures for the second part of the study

#### 2.2.2. Data set

The database created in the first part of the study containing the reference measurement and 12 months of follow-up was the starting point. But then, in order to obtain sets with varying observation lengths, the number of months was successively reduced to 8, 4, 2, and 1. In order to preserve the original nature of the changes, measurements were removed not just from the end of the period, but also from selected months, thus maintaining the trend structure. This resulted in 5 separate datasets.

To facilitate the analysis of results, including determining the threshold, the database was structured such that each difference value (0 to 39 dB) was represented by 25 cases (40 value x 25 cases = 1000 cases). This approach allowed the recommendations to be analyzed in the context of different levels of change.

#### 2.2.3. Provision of the dataset to ChatGPT

Data were provided to ChatGPT in the same way as in the first part of the study. The instructions remained consistent, with the only variation being the duration of hearing monitoring via the app, which differed in terms of the number of months.

### 2.3. Statistical analysis

All analyses were made in Matlab (version 2023b, MathWorks, Natick, MA). Percent agreement and Cohen’s Kappa [46] were used to evaluate the correctness of responses (i.e. accuracy in comparison to the experts’ score). A McNemar test [47] was used to compare the accuracy of the different approaches. In all analyses, a 95% confidence level (*p*< 0.05) was taken as the criterion of significance. Receiver operating characteristics (ROCs) and areas under the ROC curve (AUCs) were used to deduce which hearing threshold ChatGPT used to decide whether a patient should visit a specialist. This was estimated by taking the point on the ROC curve which gave the highest AUC. Fleiss’s Kappa was used to evaluate consistency [48]. The values of Kappa can be interpreted as follows: <0.0, poor; 0.01–0.2, slight; 0.21–0.4, fair; 0.41–0.6, moderate; 0.61–0.8, substantial; and 0.81–1.0, almost perfect agreement [49].

## 3. Results

### 3.1. First part – Comparison of ChatGPT decisions with experts

In this aspect, we compared how well the classifications made by ChatGPT matched those made by human experts. Could ChatGPT be used to look at a set of longitudinal observations and accurately decide when consultation with a professional is needed? Here, we utilized a dataset covering a 12-month observation period.

Five experts, along with ChatGPT (queried 5 times), rated 1,000 cases featuring different patterns of change illustrated in Figure 3. First, we evaluated consistency (Table 1). For both the experts and ChatGPT, consistency as measured by Fleiss’s Kappa was above 0.6, which is categorized as substantial agreement.

**Table 1.** Consistency (Fleiss’s Kappa) between 5 experts and 5 trials of ChatGPT.

|  | Fleiss's Kappa | *p*-level |
| --- | --- | --- |
| Experts | 0.63 | <0.001 |
| ChatGPT | 0.64 | <0.001 |

Next, we assessed the accuracy of ChatGPT. The consensus decision of the experts was defined to be when 3 or more of them agreed. Table 2 presents the accuracy and Cohen’s Kappa for each trial of ChatGPT and for the consensus decision across the 5 trials. Here, we employed as a benchmark the simple criterion of a 10 dB change – the maximum change between measurements over the 12-month period [43–45].

**Table 2.** Accuracy and Cohen’s Kappa of Δ10 dB criterion and decisions of ChatGPT in comparison to the consensus of 5 experts (≥3 of 5 experts).

| | Accuracy | Cohen's Kappa | *p*-level<br>Cohen's Kappa | ChatGPT vs Criterion of $\Delta 10$ dB |
| --- | --- | --- | --- | --- |
| ChatGPT Trial 1 | 0.80 | 0.56 | <0.001 | $\chi^2 = 0.6, p = 0.42$ |
| ChatGPT Trial 2 | 0.83 | 0.60 | <0.001 | $\chi^2 = 2.3, p = 0.12$ |
| ChatGPT Trial 3 | 0.82 | 0.59 | <0.001 | $\chi^2 = 0.02, p = 0.87$ |
| ChatGPT Trial 4 | 0.82 | 0.59 | <0.001 | $\chi^2 = 0.02, p = 0.88$ |
| ChatGPT Trial 5 | 0.84 | 0.64 | <0.001 | $\chi^2 = 3.3, p = 0.07$ |
| ChatGPT $\geq 3$ of 5 trials | 0.87 | 0.69 | <0.001 | $\chi^2 = 58.0, p < 0.001$ |
| Criterion of $\Delta 10$ dB | 0.81 | 0.54 | <0.001 | |

A single query of ChatGPT showed a level of agreement with the experts similar to that of the strict Δ10 dB HL criterion (a comparison using McNemar’s test yielded no significant difference). However, when ChatGPT was utilized multiple times, its agreement with the experts was significantly higher than when the Δ10 dB criterion was used. Therefore, in further analysis, we looked at classifications involving the most frequent response across 5 trials by ChatGPT.

To delve deeper into ChatGPT’s decision-making, we compared cases where all 5 experts agreed with those where at least one expert disagreed (Table 3). For those cases where all experts agreed, ChatGPT’s accuracy was almost identical to theirs. In instances where the experts had divided opinions, ChatGPT’s agreement was only slight (bordering on fair). In both scenarios, however, ChatGPT surpassed the simple Δ10 dB criterion in terms of accuracy relative to the average expert opinion.

**Table 3.**
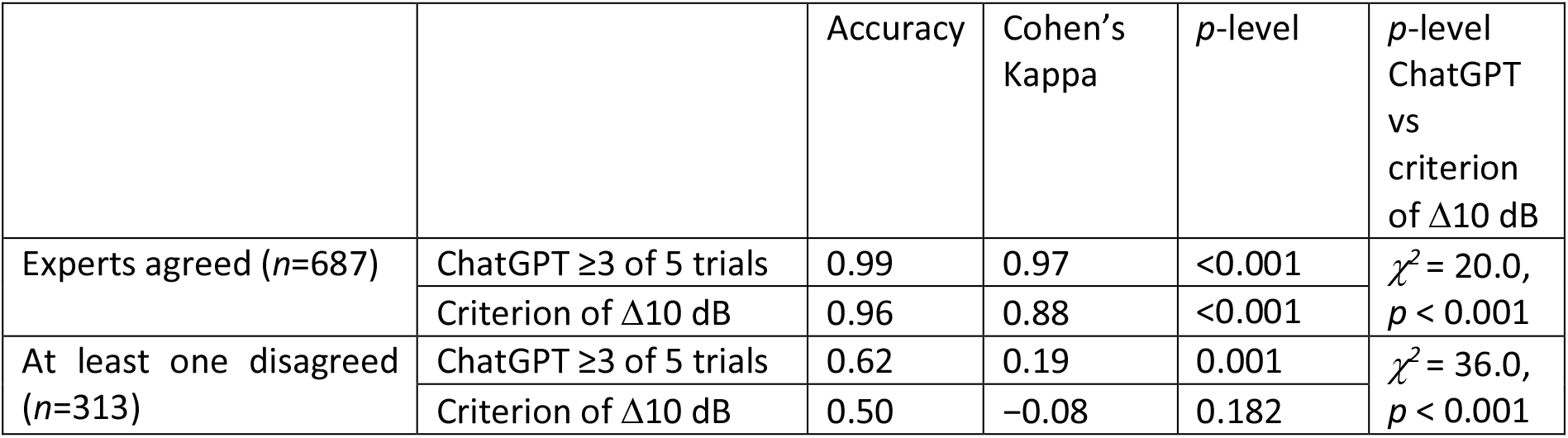
Accuracy and Cohen’s Kappa when all 5 experts agreed (top 2 rows) and when one or more experts disagreed (lower 2 rows). Comparisons are also made to when a strict criterion of Δ10 dB was used.

| | | Accuracy | Cohen's Kappa | *p*-level | *p*-level ChatGPT vs criterion of $\Delta 10$ dB |
| --- | --- | --- | --- | --- | --- |
| Experts agreed (*n*=687) | ChatGPT $\geq 3$ of 5 trials | 0.99 | 0.97 | <0.001 | $\chi^2 = 20.0$,<br>$p < 0.001$ |
| | Criterion of $\Delta 10$ dB | 0.96 | 0.88 | <0.001 | |
| At least one disagreed (*n*=313) | ChatGPT $\geq 3$ of 5 trials | 0.62 | 0.19 | 0.001 | $\chi^2 = 36.0$,<br>$p < 0.001$ |
| | Criterion of $\Delta 10$ dB | 0.50 | -0.08 | 0.182 | |

We also examined how ChatGPT handled different types of changes in longitudinal observations. In Table 4, the dataset reflects the 5 classes as shown in Figure 3: Progressive, Fluctuating, Sudden change with return to baseline, Sudden change without return to baseline, and No change. For ‘No change’ cases, the ratings from both the Δ10 dB criterion and ChatGPT matched those of the experts. Notably, ChatGPT performed significantly better than the simple criterion in the Progressive cases and in both types of Sudden change cases (regardless of whether there was a return to baseline).

**Table 4.** Accuracy and Cohen’s Kappa when the dataset was divided into different types of changes. ChatGPT data is for ≥3 of 5. For comparison, results are also shown for when a strict criterion of Δ10 dB HL was used.

| Type of change | Method | Accuracy | Cohen's Kappa | *p*-level<br>Cohen's Kappa | ChatGPT vs<br>Criterion<br>of $\Delta 10$ dB |
| --- | --- | --- | --- | --- | --- |
| Progressive (*n*=350) | ChatGPT | 0.89 | 0.08 | 0.55 | $\chi^2 = 7.1$,<br>$p = 0.007$ |
| | $\Delta 10$ dB criterion | 0.86 | 0.07 | 0.59 | |
| Fluctuating (*n*=175) | ChatGPT | 0.90 | 0.64 | 0.00 | - |
| | $\Delta 10$ dB criterion | 0.90 | 0.64 | 0.00 | |
| Sudden change and back to baseline (*n*=175) | ChatGPT | 0.66 | 0.38 | <0.001 | $\chi^2 = 43.0$,<br>$p < 0.001$ |
| | $\Delta 10$ dB criterion | 0.40 | 0.11 | 0.05 | |
| Sudden change without return to baseline (*n*=175) | ChatGPT | 0.95 | 0.66 | <0.001 | $\chi^2 = 4.2$,<br>$p = 0.04$ |
| | $\Delta 10$ dB criterion | 0.91 | 0.53 | <0.001 | |
| No change (*n*=125) | ChatGPT | 1.00 | - | - | - |
| | $\Delta 10$ dB criterion | 1.00 | - | - | |

The full responses of ChatGPT and the experts are included in the supplementary material.

### 3.2. Second part – Determination of threshold for decisions

To investigate whether ChatGPT adjusts its decision threshold based on the length of observation – expressed here by the number of observations – we examined the results of its categorization. ChatGPT was asked to categorize the data in two ways: either the subject did not need to consult (pass) or needed to consult (refer) with a specialist, based on the set of 1, 2, 4, 8, or 12 observations provided. The results for 1,000 cases at each observation period are presented in Figure 5A–E, which displays histograms of the largest differences between observations, colored according to ChatGPT’s decisions.

**Figure 5.**
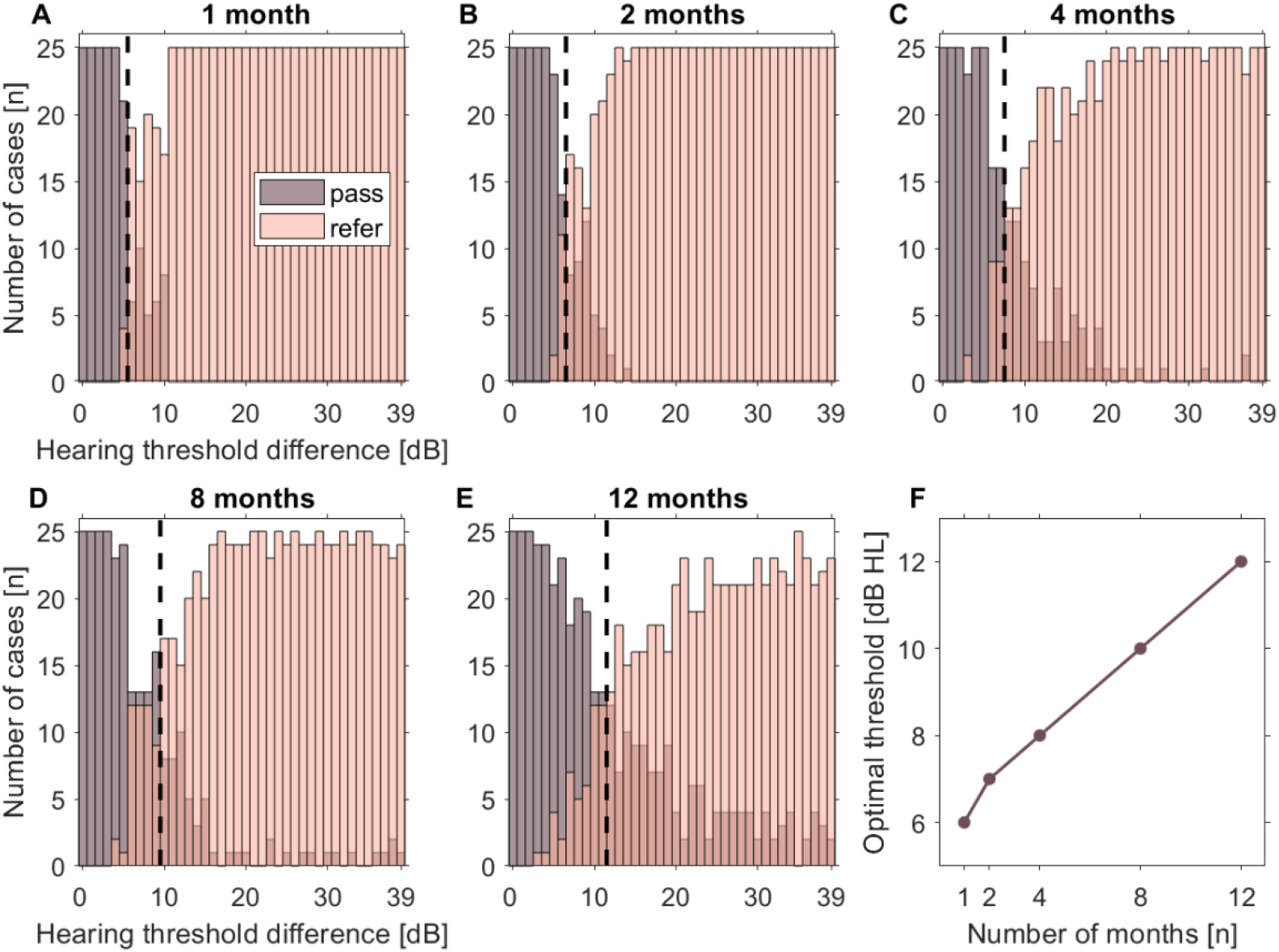
A–E: Histograms of pass/refer results of hearing thresholds rated by ChatGPT. For each 1 dB interval, there are 25 cases. In A–E, the X-axis shows the maximum PTA difference in dB, while the Y-axis represents the number of cases. The dashed lines in the A-E histograms indicate the point of best discrimination between pass and refer. F: Optimal hearing threshold values (change in hearing threshold required for a refer result) based on fitting AUC. In panel F, the X-axis represents the observation period, expressed in terms of number of monthly observations in a year, and the Y-axis shows the threshold (in dB) at which cases were referred to a specialist. The points correspond to the dashed lines from the A-E histograms.

From Figure 5A-E, it is evident that the cross-over point between cases classified as pass and refer shifts to longer values as the observation period increases. Additionally, with more observations available, there are more cases exhibiting significant differences during the observation period that are still classified as pass. A ROC analysis was performed on this dataset to determine the threshold values that yield the highest AUC (i.e. best possible discrimination between pass and refer); these thresholds are provided in Figure 5F. Notably, the discrimination level – the change in hearing threshold that most effectively separates the pass and refer cases – increased from Δ6 dB for 12 1-monthly observations to Δ12 dB for a single observation over the 12 months.

These findings indicate that ChatGPT does not apply a single criterion for all observation periods. Instead, it makes use of all the available data to make more nuanced decisions. ChatGPT’s responses for all the months of observations are available in the supplementary material.

## 4. Discussion

In this study, we explored the potential of combining two existing technologies – mobile hearing-test apps and ChatGPT – to maximize the utility of these tools. While our tests are not definitive, they demonstrate ChatGPT’s promise as a flexible tool for hearing monitoring. Its ability to adjust decision thresholds based on observation length, which aligned closely with the consensus of the experts, highlights its effectiveness, particularly in straightforward cases. A strategy of asking ChatGPT the same question multiple times further improved accuracy and reinforced reliability.

ChatGPT also showed advantages over rigid algorithms, offering a more nuanced analysis of complex audiometric data and demonstrating a strong potential for long-term monitoring. With more observations available, there are more cases exhibiting significant differences during the observation period that are still classified as pass, as ChatGPT recognizes single, non-consequential changes and avoids unnecessarily referring such cases to specialists. This contrasts with shorter observation periods, where a single change may appear more concerning. In less complex scenarios, it achieved near-perfect agreement with experts. Although its performance was lower in cases with divided expert opinions, ChatGPT still surpassed a strict decision criterion. However, this highlights the need for further refinement, and expert involvement remains essential for managing more complex scenarios. Furthermore, the analysis of different types of hearing loss revealed that ChatGPT was particularly effective in identifying progressive and abrupt changes, showcasing its utility in interpreting dynamic clinical patterns.

Our findings sit within the broader context of bringing AI to the field of medicine. Research to date has shown that models such as ChatGPT can effectively support patient triage [50] and making diagnoses based on case descriptions [51]. AI’s capacity to analyze large datasets and discern pertinent trends, as identified by other researchers [52], underscores the prospect of it being used in the long-term monitoring of patients.

AI tools have the potential to enhance communication between patients and physicians [53]. By answering patients’ questions, they can provide additional information, increasing patients’ awareness of their health status, and encouraging them to undertake regular monitoring. Integrating AI with mobile apps could be a further step towards personalized health monitoring [54]. Real-time data analysis has the potential to quickly identify worrying trends and help with making accurate decisions, both in the hands of patients and healthcare professionals. Such assistance would be welcome in regions with limited access to specialist medical care.

Despite the promising prospects, it is also important to highlight the limitations of this technology. While AI can speed up analysis and facilitate decision-making [55], ChatGPT is less effective in more complex clinical cases that require weighing up of wider medical issues. As Marshall [56] observed, LLMs are capable of making a differential diagnosis from limited input data, but in more difficult cases a specialist can use clinical knowledge and experience, as well as skill in interviewing and performing physical examinations. For example, when recommending changes in drug treatment [57], or proposals for additional specialist examinations [58], AI models tend to recommend an excessive number of follow-ups. Thus, the responses given by AI should be regarded as preliminary indications, and they require validation by an expert to prevent unnecessary interventions and expense.

In summary, in an audiological setting, ChatGPT shows considerable promise as a tool for enhancing self-monitoring. It can provide recommendations for action based on on-going hearing tests. Patients might be able to use ChatGPT to get preliminary guidance on whether a specialist consultation is warranted, while clinicians can leverage its ability to efficiently manage larger patient populations. By delegating routine tasks to AI, a healthcare professional could allocate more time to complex cases, improving overall patient care. The integration of AI-driven decision-making with hearing monitoring tools appears imminent. Until then, this combination of existing technologies offers an accessible and practical interim solution for both patients and healthcare providers.

## 5. Conclusions

This study underscores the potential of ChatGPT and similar chatbots based on LLMs as tools to support hearing monitoring. By achieving high agreement with expert assessments, particularly through the use of a multiple-response strategy, ChatGPT can provide valuable support in long-term monitoring. Linked to a mobile hearing app, it can identify problems at an early stage and suggest an appropriate action. However, ChatGPT’s role needs to remain supportive, complementing rather than replacing clinical expertise, especially in complex scenarios. Continued advancements in AI are expected to further enhance its capabilities, making health monitoring more efficient and accessible, particularly in cases involving long data sets.

## Data Availability

All data produced in the present work are contained in the manuscript and in the supplementary material

## 6. Acknowledgment

The authors thank also Andrew Bell for comments on an earlier version of this manuscript.

## Notes

### Competing Interest Statement

The authors have declared no competing interest.

### Funding Statement

This study did not receive any funding

